# Retrospective study of botulinum neurotoxin for hemifacial spasm: experience over 25 years

**DOI:** 10.1101/2023.10.03.23296410

**Authors:** Moon Keen Lee

## Abstract

**Introduction:** Hemifacial spasm (HFS) is a not uncommon, debilitating movement disorder. This retrospective study examines 25 years of experience with Botulinum neurotoxin (BoNT) injections for HFS in Malaysia.

**Objective:** To assess BoNT’s long-term effectiveness in treating HFS in a private practice.

**Methods:** Data from 35 patients, including demographics, treatment outcomes, and complications, were collected from a private clinic over 25 years.

**Results:** BoNT demonstrated a decreasing dose trend over time, with extended treatment intervals. Complications were rare and transient, with high patient retention.

**Conclusion:** BoNT is a safe and effective long-term HFS treatment, enhancing accessibility in private practice, complementing public healthcare services.

## INTRODUCTION

Hemifacial spasm (HFS) is a movement disorder in which there is involuntary twitching of the eye and face on one side. It affects 14.5 per 100,000 women and 7.4 per 100,000 men. Although rare, it often persists for years, causing considerable disability.

Chemodenervation with botulinum neurotoxin (BoNT) injections was pioneered by Scott et al in the 1970’s for treatment of eye conditions^1^ and was approved by the FDA in 1989 for blepharospasm and strabismus in adults.

BoNT is now widely used for movement disorders in various parts of the body such as dystonia and spasticity. Its use been described in Malaysia for limb spasticity in children^2^ but treatment for eye conditions has not been reported. In the author’s practice, the main indications for BoNT are HFS and blepharospasm, both conditions which cause involuntary twitching of the eyes and face.

Despite medication advances over the past two decades, BoNT is still the best short and medium term treatment for these disorders.

This study describes the results of BoNT injection for HFS in a private practice in Malaysia.

## OBJECTIVE

A retrospective study of treatment outcomes of BoNT in adults for HFS was conducted on a series of patients in a private practice setting in Malaysia.

## METHODS

Data were compiled from consecutive patients who were treated with BoNT at Pantai Hospital Kuala Lumpur and Alpha Specialist Centre over the period January 1998 - August 2023. Data included age at diagnosis, duration of illness, duration of follow-up, dose of BoNT, mean interval between injections, and treatment and complication outcomes. Mean interval between injections was calculated as the average of five of the most recent consecutive treatment intervals. Prolonged intervals caused by the pandemic lockdown from March 2020 to March 2022 were not included. All patients had failed medical therapy before opting for BoNT.

Pre-existing facial weakness in HFS patients was documented and its progress was followed up.

Where feasible, a vial of BoNT was co-shared by patients in order to reduce cost. Formulations used were either Botox® or Dysport®. In order to rationalize reporting, units of Dysport® were converted to Botox® units by a 3:1 factor.^3^ The areas injected were as illustrated in Figure 1.^4^

**Figure 1.**
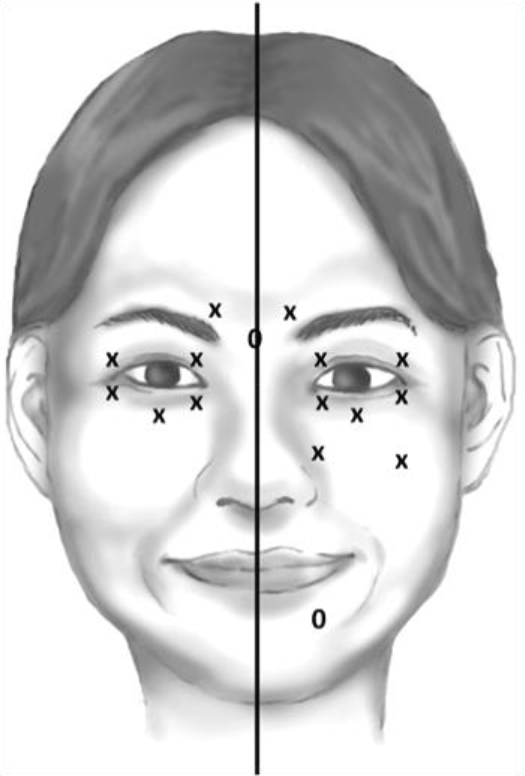
The injection sites scheme for the treatment of hemifacial spasm (HFS, right part). The dose was 2.5 units per point. 6-7 periocular points and 2-3 facial points on affected site are injected. ‘x’ means fixed injection point and ‘o’ means elective injection point. – After Reference 4.

The author’s practice is to deliver about 80% of the estimated total dose and to top up after two weeks as needed. Injecting around the mouth area was avoided as far as possible, to prevent drooping of the mouth.

For the purpose of comparison, the most recent five treatments were computed, in order to control for changes in BoNT formulation and evolution of injection techniques.

## RESULTS

### Diagnosis of Patients treated with BoNT (Table 1)

|  |  |
| --- | --- |
| Hemifacial spasm | 35 |
| Blepharospasm | 8 |
| Dystonia | 8 |
| Synkinesis | 4 |
| Spasticity | 1 |
| Miscellaneous | 1 |
| No Trace | 11 |
| **TOTAL NO. OF PATIENTS** | **68** |

### Hemifacial Spasm: Patient Characteristics (Table 2)

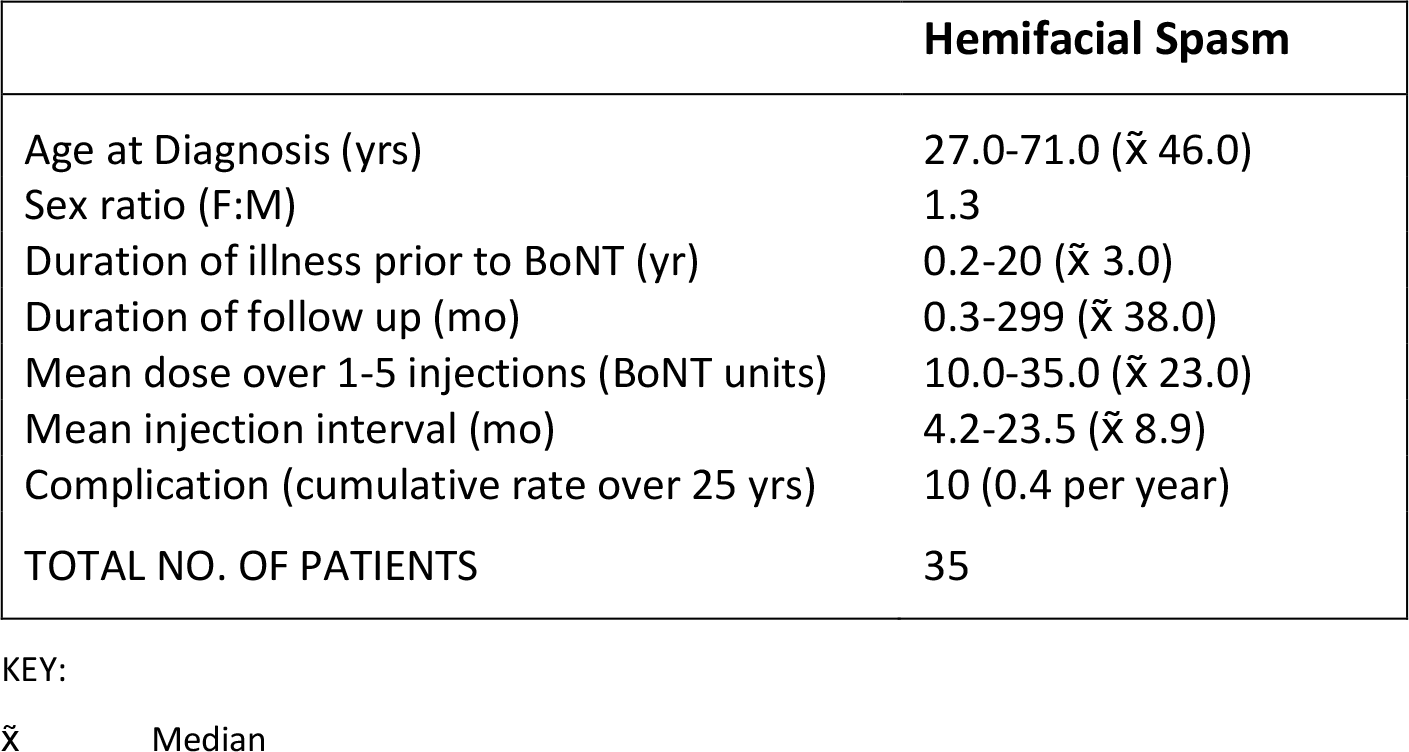

A total of 68 patients were treated with BoNT over the period January 1998 - August 2023.

There were 36 cases of HFS (one record not retrieved). The median age at presentation was 46 years, with female preponderance of 1.3. Median duration of illness prior to BoNT was quite prolonged at 3 years. There was a slight predilection for the left side at 57%. Total number of treatments ranged from 1 to >10 over a span of 25 years. Mean treatment interval ranged from 4.2 to 23.5 months (median 8.9 months).

There was a trend towards lower BoNT doses between Injections 1 and 5: down from 23.3 to 20.0 units (Figure 1).

**Figure 1:**
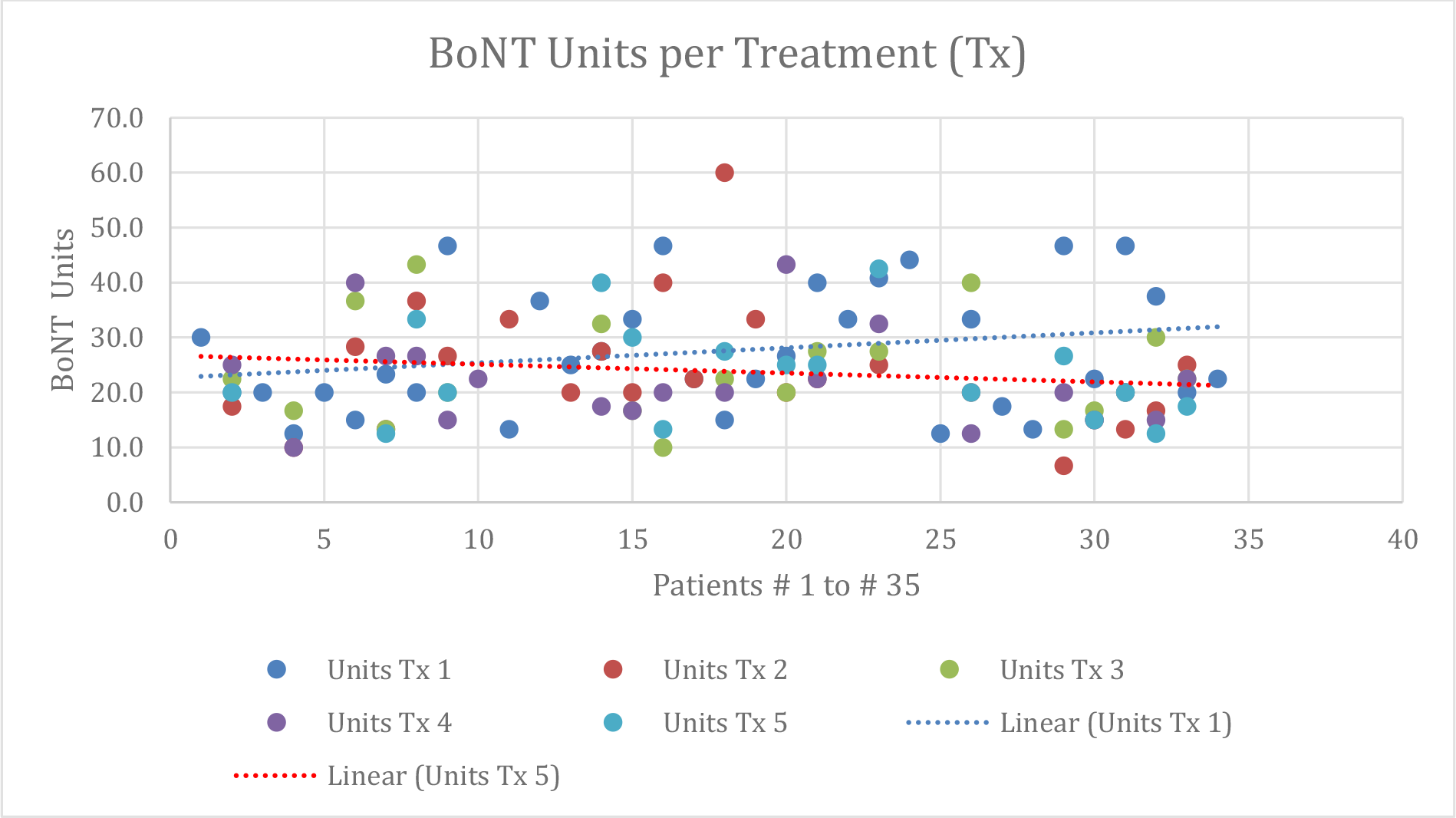
Hemifacial Spasm: Trend Towards Decrease in BoNT Doses Over Time KEY: Units Tx 1…5 - Units given during Treatment 1…5

Intervals between Treatments 4 and 5 were longer than Intervals between Treatments 1 and 2, up from median of 8.0 to 10.5 months, suggesting amelioration of severity with continued treatment (Figure 2).

**Figure 2:**
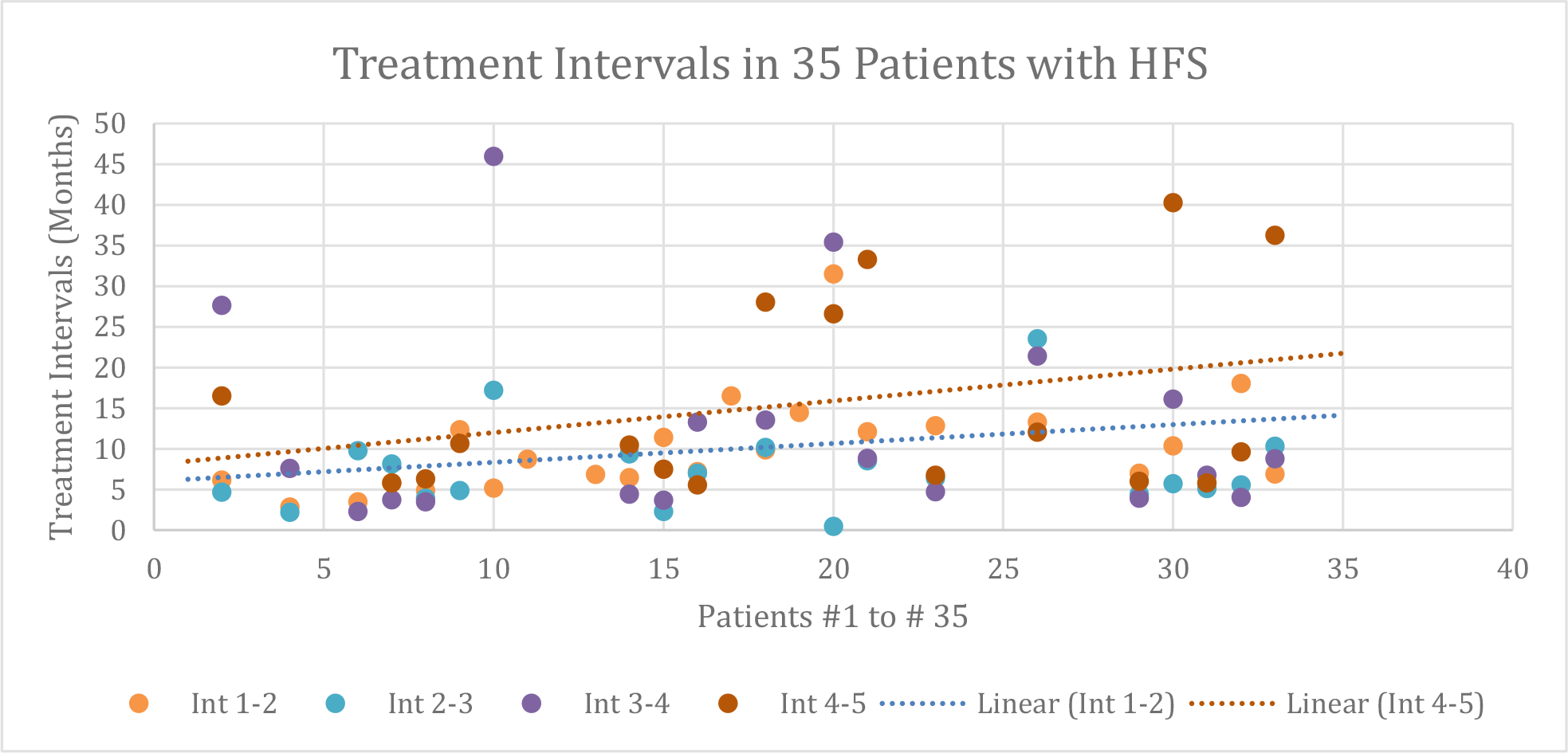
Hemifacial Spasm: Trend towards Increase in Treatment Intervals Over Time KEY (time interval between consecutive treatments (months): Int 1-2 Interval between Treatments 1 and 2 Int 2-3 Interval between Treatments 2 and 3 Int 3-4 Interval between Treatments 3 and 4 Int 4-5 Interval between Treatments 4 and 5

Complication rate in HFS patients was 10 incidents over 25 years of treatment, i.e. 0.4 per year. These consisted of ptosis or drooping of the mouth. All adverse effects were transient and resolved within one month. One patient did not pursue further treatment as a consequence of complication.

No patient opted for subsequent neurosurgery.

## DISCUSSION

This study presents the results of BoNT for hemifacial spasm in a private practice setting in Malaysia. The patient profile and results are comparable with other reports in the literature.

HFS symptoms preceded BoNT treatment for a median duration of 3 years. This was related to searching for the correct diagnosis as well as trial of various medications.

There were 10 incidences of transient drooping of the eyelid or angle of the mouth over 25 years. This is likely to be caused by pre-existing facial weakness in these patients.

No complication of infection occurred in any patient.

Overall patient retention was good, with only one dropout from treatment. No patient had to resort to neurosurgery.

The effect of BoNT typically lasts for about three months. In this study, our patients managed their condition with median treatment intervals of 9 months. This resulted in amortized BoNT treatment cost approaching that of medication.

Wait times for BoNT are long in public hospitals in Malaysia. Rising income levels, combined with cost-sharing of vials, has rendered BoNT affordable in private practice. This significant improvement in quality of life is thereby available to middle income patients, to complement public hospital services. BoNT is now within reach for most urban or suburban patients in Malaysia, whether in the public or private health sector. As for the non-urban population, those patients will have to be served through injection out-reach efforts.

## CONCLUSION

BoNT is confirmed to be a safe and effective treatment for hemifacial spasm on long term follow up over 25 years. BoNT doses tended to decrease and treatment intervals to extend upon continuous treatment. This finding, coupled with cost-sharing of BoNT among patients, has enabled private practice to supplement resource-strapped public hospital services in Malaysia.

## Data Availability

All data produced in the present study are available upon reasonable request to the authors

